# Efficacy of Highly Aspherical Lenslet(HAL) spectacles in slowing myopia progression in children and adolescents: A multi-centre, retrospective, real-world study in India-SOLIDITY study

**DOI:** 10.64898/2026.04.10.26350578

**Authors:** Rohit Saxena, Jitendra Jethani, Lipika Roy, Jyoti Matalia, Pavan Kumar Verkicharla, Sandra Ganesh, Akshya Parthasarathy, Suraj Nayak, Vinay Gupta, Kalpana Narendran, Pratibha Panmand, Prajna Ghosh, Sumitha Muthu, Kunal Srivastava, Olga Prenat

## Abstract

**Objective:** The study aims to evaluate the real-world effectiveness of Highly Aspherical Lenslets spectacle (HAL; Essilor^®^ Stellest^®^) in slowing myopia progression among Indian children and adolescents aged between 4 and 16 years.

**Methods and analysis:** This was a multicentre retrospective study conducted across 10 leading ophthalmic centers. The study participants comprised children aged between 4 and 16 years who were prescribed HAL spectacles (Essilor^®^ Stellest^®^). Data were extracted from electronic medical records at three time points: T1: One year prior to intervention; T2: Baseline at HAL spectacle prescription; T3: 6 to 24 months after prescription. The primary endpoint was the myopia progression and axial elongation in the year following prescription, compared with the untreated year and with published meta-regression models. Only data from the right eye were analysed, with the expected physiological progression estimated based on the individual progression trajectory after adjusting for age-related slowing as reported in published meta-regression models.

**Results:** A total of 372 myopic children were included in the study. The annual myopia progression was –0.72 ± 0.47 D/year during the untreated period, reducing to –0.11 ± 0.29 D/year with HAL spectacle wear. The expected progression without treatment was –0.65 D/year, based on trajectory-adjusted modelling, indicating a treatment effect of 0.54 D/years and an estimated 83% slowing in myopia progression compared to expected progression. The expected axial elongation under physiological conditions was 0.29 mm/year, estimated using age-adjusted meta-regression models; with HAL lens wear, axial elongation was 0.11 ± 0.16 mm/year, corresponding to a ∼62% relative slowing of elongation.

**Conclusion:** The present study demonstrates the real-world evidence validating the efficacy of HAL lenses as an effective myopia control intervention in Indian children and adolescents. The retrospective design and limited follow-up period warrant future prospective, long-term studies to validate these findings.

## INTRODUCTION

Myopia has become a global public health concern, recognized by the World Health Organization (WHO) as an immediate priority leading to visual impairment.[1] By 2050, myopia is estimated to affect ∼50% of the population worldwide, with 10% manifesting as high myopia.[2] Increasing levels of myopia elevate the risk of serious ocular complications, such as myopic maculopathy, retinal detachment, and glaucoma.[3] Additionally, myopia results in uncorrectable visual impairment, imposes a substantial economic burden on affected individuals, and places considerable strain on the public healthcare system.[4] While prevalence varies with geographic region, myopia remains a predominant cause of visual impairment worldwide, affecting populations in both developing and developed nations.[5,6]

The overall crude prevalence of myopia in Indian children aged 5 to 15 years over four decades (1980-2020) in a meta-analysis was reported as 7.5%.[7] The prevalence of myopia was higher among urban children (8.5%; 95% CI: 7.1–9.9%) compared with rural children (6.1%; 95% CI: 4.5–7.7%). The highest prevalence was observed in urban children aged 11–15 years, reaching 15.0% in the most recent decade. The rise in childhood myopia in India is potentially due to prolonged near work time, reduced outdoor activities, and increased academic pressure.[8] Factors influencing myopia prevalence in Indian children include age, educational status, gender, and parental history of myopia.[9–11]

Myopia typically develops during the early school years [12] and progresses as it increases with age [13]. A retrospective Indian study conducted between January 2010 and January 2016 included participants with myopic refractive error, aged 1–30 years. Mean myopia progression varied across age groups, ranging from −0.07 ± 0.02 D to −0.51 ± 0.02 D, with the fastest progression observed in children aged 6–10 years and the slowest in the 26–30 years age group. Age-stratified analysis found that participants younger than 15 years showed significantly greater myopia progression (−0.45 ± 0.01 D) compared with those older than 15 years (−0.14 ± 0.01 D; p < 0.001).[14] Since myopia progresses more rapidly in children, early identification and timely intervention are crucial.

Slowing myopia progression even by 1 D during childhood may reduce lifetime myopic maculopathy risk by ∼40%.[15] Consequently, interventions to slow myopia progression (myopia control) have gained enormous attention among the paediatric population. Optical and pharmacological interventions such as low-dose atropine, orthokeratology, and multifocal contact lenses are effective in slowing myopia progression.[16] However, adoption of these options into routine clinical practice is limited by the long-term efficacy and safety concerns of newer modalities.[17,18] Despite the growing need to effectively treat myopia, challenges related to affordability, acceptability, long-term adherence, and continuity of care remain unaddressed. This highlights the importance of identifying and implementing myopia control strategies tailored to the Indian population.

Spectacles are widely accepted as a non-invasive, feasible, and easily wearable optical intervention, making them a suitable option for use in children.[19,20] Recent advances in spectacle lens designs have moved myopia management forward.[21,22] The Essilor^®^ Stellest^®^ lenses launched in 2020 incorporate Highly Aspherical Lenslet Target (H.AL.T.) technology, comprising 1,021 highly aspherical contiguous lenslets arranged in 11 concentric rings. This optical design generates a volume of myopic defocus (VoMD) in front of the retina, thereby slowing myopia progression.[23,24] A two-year randomized clinical trial showed that a highly aspherical lenslet (HAL) spectacle reduced myopia progression by 0.80 D and axial elongation by 0.35 mm.[22] Additionally, the five-year follow-up showed that the use of HAL delayed myopia progression and axial elongation by three years compared with single-vision lenses (SVL).[25]

The real-world effectiveness of HAL in slowing myopia progression in the Indian population remains unexplored. Thus, the present multicenter, retrospective study aims to investigate the effectiveness of HAL lenses in slowing myopia progression in Indian children and adolescents aged 4 to 16 years.

## METHODS

### Study Design

The SOLIDITY study is a multi-centre retrospective study evaluating the real-world effectiveness of HAL lens spectacles in slowing myopia progression among Indian children and adolescents. Data was collected from 10 leading ophthalmic centers between July 2022 and June 2024. The study adhered to the Declaration of Helsinki. Ethical approval was obtained from the ACE Independent Ethics Committee and subsequently from institutional review boards at all participating centres. Due to the retrospective and anonymised nature of the data, individual informed consent was waived.

### Participants

The study included children aged 4–16 years with documented progressive myopia who were prescribed HAL lenses (Essilor® Stellest®) during the study period. All participants had been managed with SVL during the pretreatment period. Progressive myopia was defined as an increase in spherical equivalent refraction (SER) of at least −0.25 D within the 12 months preceding HAL lens prescription. A subgroup of patients received concomitant atropine therapy. In all such cases, atropine 0.01% had been initiated prior to HAL lens prescription while the patients were wearing SVL and was continued during HAL lens wear. This treatment pathway reflects common real-world clinical practice in India, where children are typically started on atropine 0.01% with SVL, and subsequently escalated to myopia control spectacle lenses if progression continues.

The participants’ follow-up period was standardized to 12 ± 1 months to ensure comparability of outcomes. Comparison of unimputed (n = 217) and imputed (n = 372) datasets for SER and axial length showed no statistically significant differences at any time point (all p >0.05) (S1 Table and S1 Figure), thereby confirming that the findings are robust and not influenced by data imputation.

### Study Procedures

Anonymised data were extracted from electronic medical records, optical dispensing logs, and investigation reports for each study centre. Demographic variables included age, sex, and clinical history. SER and axial length were captured at three time points: T1 = One year prior to intervention, T2 = Baseline at HAL prescription, and T3 = 1-year post HAL lens wear.

Cycloplegic refraction and axial length measurements were performed at all participating Myopia Excellence Centres as part of standard care. Axial length was measured using non-contact partial coherence interferometry. Measurements were performed by trained optometrists or ophthalmologists using standardised protocols and calibrated equipment at each study interval. Due to the retrospective nature of the study, the specific models of autorefractors and biometers were not standardized or uniformly documented, and measurements were obtained using validated instruments available at each center. The primary outcomes were myopia progression and axial elongation. Evaluated covariates were age (≤10 years vs. >10 years) and the adjunct use of atropine in combination with HAL lenses (with atropine and without atropine).

### Statistical Analyses

Statistical analysis was carried out on both the unimputed dataset (n = 217) and the full dataset after multiple imputation (n = 372). Values that fell outside the protocol-specified 11–13-month follow-up window were treated as missing and were imputed for the purpose of analysis because they did not represent valid 12-month outcomes. The out-of-window measurements (12± 1 months) were imputed using Multiple Imputation by Chained Equations (MICE). MICE was chosen because it preserves the multivariable structure of the data and generates multiple plausible estimates for the intended 12-month value by using information from baseline characteristics and longitudinal covariates. Twenty imputed datasets were created and pooled to minimize bias and to maintain the underlying distribution of key variables such as SER and axial length. The use of MICE prevented misclassification of out-of-window values as true 12-month measurements, avoided loss of sample size, and reduced the risk of biased mean estimates that can occur when irregularly timed observations are used without adjustment. The process of selecting subjects and the exclusion criteria employed to determine the final analysis population for trajectory-adjusted modelling is detailed in Supplementary Figure 1.

Only data from the right eye were analysed. To ensure completeness and alignment with previously published meta-regression comparisons, both bilateral and averaged binocular analyses were conducted for trajectory-adjusted expected progression modelling.

Continuous variables were summarized as mean ± standard deviation (SD). Comparisons between unimputed and imputed datasets were assessed using independent-samples t-tests, with a two-sided p < 0.05 considered statistically significant. For longitudinal changes, observed progression during the HAL-treated year (T3–T2) was compared with trajectory-adjusted expected progression derived from individual pre-treatment progression (T2–T1), adjusted for age-related slowing using published meta-regression models [26,27]. Treatment effects were further benchmarked against modelled expectations, incorporating approximately 9.7% annual slowing of myopia progression [26] and 15% annual slowing of axial elongation [27]. Effect size (Cohen’s d) was calculated for primary outcomes, and statistical power was estimated using a paired comparison framework with a two-sided α = 0.05. Subgroup analyses were performed by age (≤10 years vs. >10 years) and by concomitant atropine use, using between-group t-tests on myopia progression and axial elongation.

## RESULTS

### Study population

A total of 372 children with myopia were included in the study (Table 1). Comparison of unimputed and imputed datasets showed no significant differences in SER or axial length measurements (Supplementary Table 1). The mean (±SD) age at the time of HAL prescription (T2) was 10.7 ± 2.6 years. A total of 173 (46.5%) participants were ≤10 years of age, and 199 (53.5%) participants were >10 years. Atropine was administered to 100 children (26.9%), while 272 children (73.1%) did not receive any adjunctive treatment for myopia.

**Table 1:** Baseline characteristics of participants.

| Parameter | n= 372 |
| --- | --- |
| Gender |  |
| Female (%) | 208 (55.9) |
| Male (%) | 164 (44.1) |
| Age at HAL Prescription (T1, years) Mean ± SD | 10.7 ± 2.6 |
| Age Category at HAL Prescription |  |
| $\leq 10$ years (%) | 173 (46.5) |
| $> 10$ years (%) | 199 (53.5) |
| Parental History of Myopia |  |
| One parent (%) | 100 (26.9) |
| Both parents (%) | 41 (11.0) |
| None (%) | 231 (62.1) |
| History of Myopia in Siblings |  |
| None (%) | 325 (87.4) |
| 1 sibling (%) | 43 (11.6) |
| 2 siblings (%) | 4 (1.1) |
| Concomitant Pharmacologic Intervention (0.01% Atropine) |  |
| None (%) | 272 (73.1) |
| 0.01% Atropine (%) | 100 (26.9) |

*Data are presented as n (%), unless stated otherwise.*
*HAL: Highly Aspherical Lenslets; SD: standard deviation*

### Myopia progression with and without HAL lenses

Table 2 presents the mean (±SD) refractive error (in SER) and myopia progression for 372 children across untreated and HAL treatment periods. During the untreated period (T2–T1), myopia progression was –0.72 D. Following the prescription of HAL lenses (T3–T2), progression was markedly reduced to –0.11 D—a slowing of 0.61 D (p = 0.001; Table 2). The SER progression in the HAL-treated year was significantly lower than the trajectory-adjusted expected progression. Based on trajectory-adjusted modelling incorporating a 9.7% annual slowing of myopia progression,[26] the expected untreated progression in the second year is −0.65 D, substantially greater than the HAL-treated value of −0.11 D. Compared to Smotherman’s trajectory-adjusted expected progression of −0.653 D, observed SER progression with HAL lenses was −0.11 D, indicating that HAL lenses reduced myopia progression by 0.545 D (83.5% reduction) (Table 3).

**Table 2:**
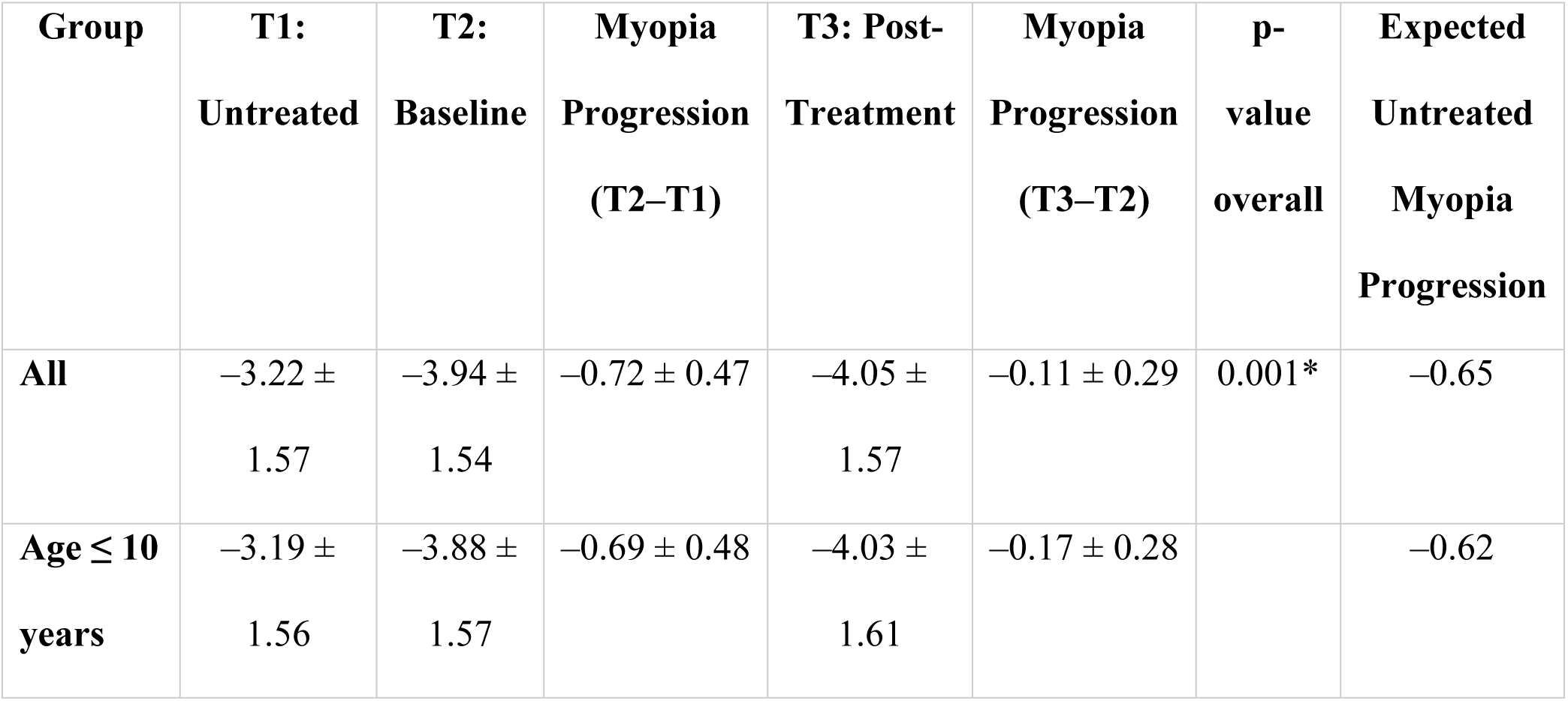
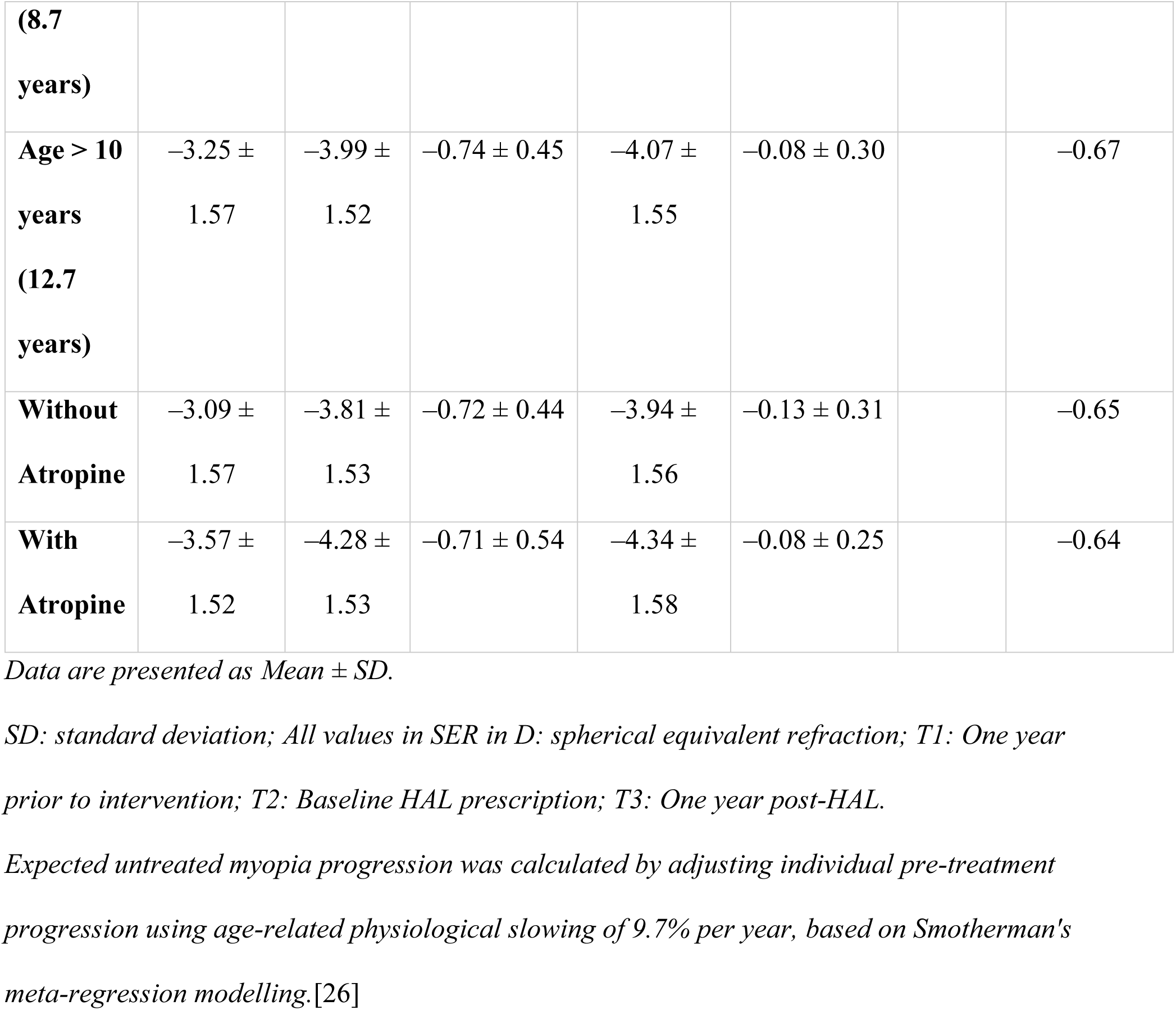
Myopia progression from the untreated year (T1) to 1-year post-HAL wear (T3)

**Table 3:** Comparative slowing of myopia progression with HAL lenses versus untreated and Smotherman’s expected progression.

| Eye<br>(N=372) | Untreated<br>$\Delta U$ (D) | HAL $\Delta H$<br>(D) | %<br>Progression<br>Control | Smotherman Expected<br>$\Delta E$ (D) | $\Delta(E-H)$ | % Lower<br>than<br>expected<br>with HAL<br>lens wear |
| --- | --- | --- | --- | --- | --- | --- |
| Right Eye | -0.718 | -0.111 | 84.5% | -0.648 | -0.537 | 82.9% |
| Left Eye | -0.728 | -0.104 | 85.7% | -0.657 | -0.553 | 84.2% |
| Both Eyes | -0.723 | -0.108 | 85.1% | -0.653 | -0.545 | 83.5% |

HAL: Highly Aspherical Lenslets; HAL year (H): annual myopia progression during HAL treatment (T3–T2); Untreated year (U): annual myopia progression prior to HAL treatment (T2–T1); Smotherman expected progression (E): calculated by adjusting untreated progression using age-related physiological slowing of 9.7% per year, based on meta-regression modelling [26]; $\Delta(E-H)$ represents the absolute difference between expected untreated progression and observed progression during HAL treatment; % Progression Control = $(1 - H/U) \times 100$ ; % Lower than expected with HAL lens wear = $(1 - H/E) \times 100$ ; Negative values indicate myopic progression.

### Axial elongation with HAL lenses

The axial elongation from baseline (T2) to 1-year post-HAL wear (T3) was 0.11 ± 0.13 mm (Table 4), which was significantly lower than the expected physiological elongation (p = 0.001). According to age-adjusted meta-regression modelling for the study cohort (mean age 10.7 years), the expected untreated axial elongation was 0.289 mm (Table 4). Compared to Brennan et al.’s expected axial elongation of 0.289 mm [27], axial elongation with HAL lenses was 0.11 mm, indicating that HAL lenses reduced axial elongation by 0.179 mm (61.9%) (p = 0.001; Table 5).

**Table 4:** Subgroup analysis of axial elongation with HAL Lenses vs. Brennan et al.’s Expected Model.

| **Group** | **T2:**<br>**Baseline**<br>**(mm)** | **T3: After 1**<br>**year HAL**<br>**wear (mm)** | **Mean axial**<br>**elongation (T3–**<br>**T2) (mm)** | **Expected**<br>**untreated axial**<br>**elongation (mm)** | **Slowing**<br>**(mm)** |
| --- | --- | --- | --- | --- | --- |
| **All** | 24.71 ±<br>1.00 | 24.82 ± 1.00 | 0.11 ± 0.13 | 0.289 | 0.179 |
| **Age ≤ 10**<br>**years** | 24.54 ±<br>0.86 | 24.68 ± 0.87 | 0.15 ± 0.16 | 0.318 | 0.168 |
| **Age > 10**<br>**years** | 24.84 ±<br>1.07 | 24.92 ± 1.08 | 0.09 ± 0.10 | 0.261 | 0.171 |
| **Without**<br>**Atropine** | 24.71 ±<br>1.02 | 24.82 ± 1.01 | 0.12 ± 0.13 | 0.289 | 0.169 |
| **With**<br>**Atropine** | 24.70 ±<br>0.92 | 24.80 ± 0.97 | 0.09 ± 0.15 | 0.289 | 0.199 |

Data are presented as Mean ± SD in millimetres (mm). Mean axial elongation represents the change in axial length between T2 and T3. HAL: Highly Aspherical Lenslets; SD: standard deviation; T2: Baseline year (HAL prescription); T3: 1-year post-HAL wear.

**Table 5:**
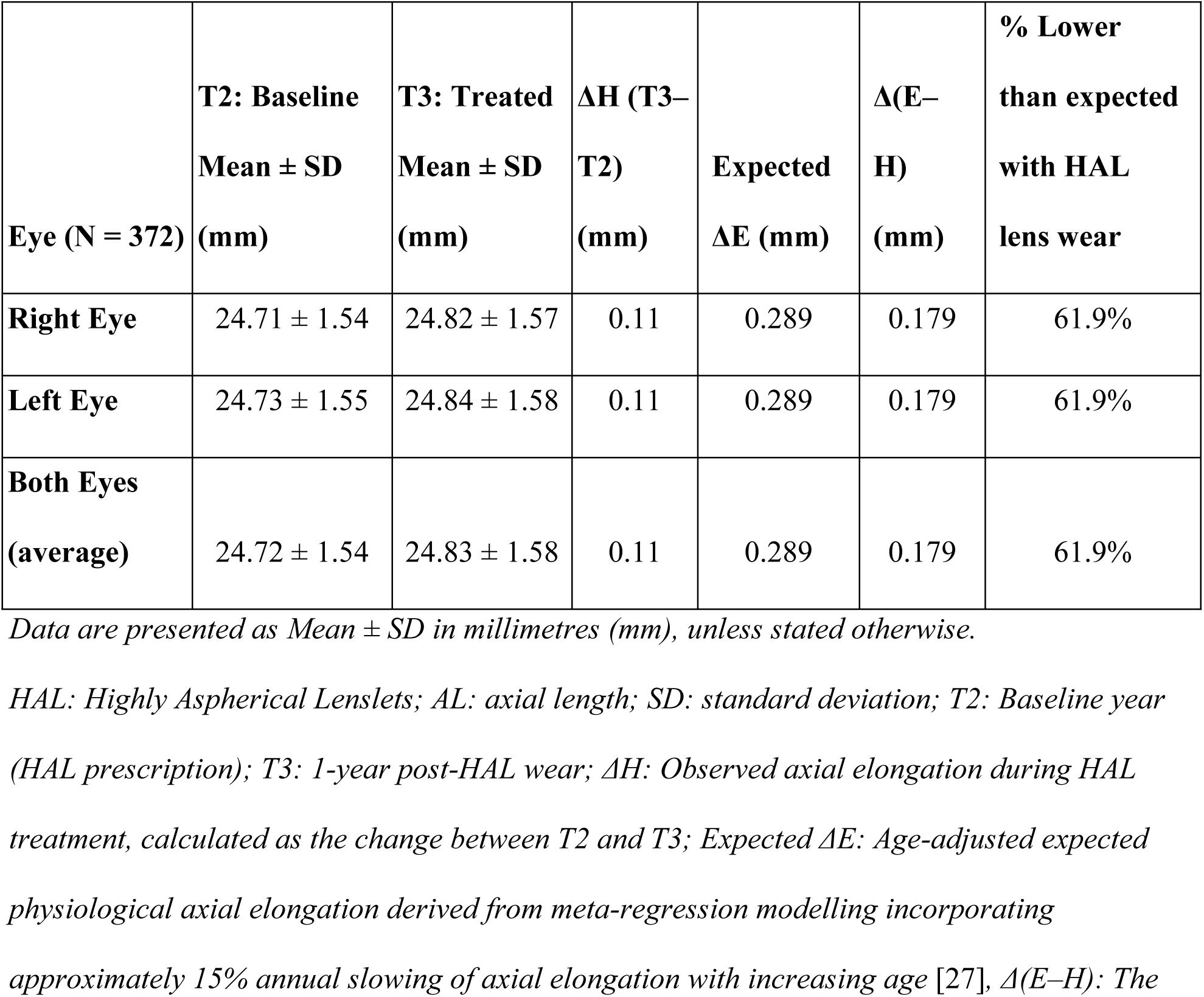
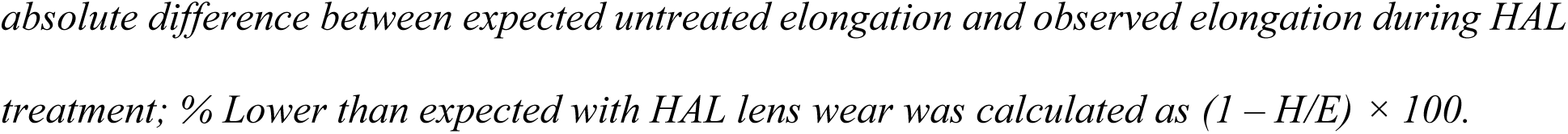
Comparative axial length (AL) progression: HAL lenses vs. Brennan et al.’s expected model.

| **Eye (N = 372)** | **T2: Baseline**<br>**Mean ± SD**<br>**(mm)** | **T3: Treated**<br>**Mean ± SD**<br>**(mm)** | **$\Delta H$ (T3–**<br>**T2)**<br>**(mm)** | **Expected**<br>**$\Delta E$ (mm)** | **$\Delta(E-H)$**<br>**(mm)** | **% Lower**<br>**than expected**<br>**with HAL**<br>**lens wear** |
| --- | --- | --- | --- | --- | --- | --- |
| **Right Eye** | 24.71 ± 1.54 | 24.82 ± 1.57 | 0.11 | 0.289 | 0.179 | 61.9% |
| **Left Eye** | 24.73 ± 1.55 | 24.84 ± 1.58 | 0.11 | 0.289 | 0.179 | 61.9% |
| **Both Eyes**<br>**(average)** | 24.72 ± 1.54 | 24.83 ± 1.58 | 0.11 | 0.289 | 0.179 | 61.9% |

Data are presented as Mean ± SD in millimetres (mm), unless stated otherwise.
HAL: Highly Aspherical Lenslets; AL: axial length; SD: standard deviation; T2: Baseline year (HAL prescription); T3: 1-year post-HAL wear; $\Delta H$ : Observed axial elongation during HAL treatment, calculated as the change between T2 and T3; Expected $\Delta E$ : Age-adjusted expected physiological axial elongation derived from meta-regression modelling incorporating approximately 15% annual slowing of axial elongation with increasing age [27], $\Delta(E-H)$ : The
*absolute difference between expected untreated elongation and observed elongation during HAL* *treatment; % Lower than expected with HAL lens wear was calculated as $(1 - H/E) \times 100$ .*

### Subgroup analysis

The slowing of myopia progression and axial elongation was statistically significant in both age groups, ≤10 years and >10 years (Tables 2 and 4), with observed progression and elongation significantly lower than the trajectory-adjusted and age-adjusted expected physiological progression (p < 0.05 for all comparisons). Similarly, significant slowing of myopia progression and axial elongation was observed in children receiving atropine and those who did not use atropine (Tables 2 and 4). In both groups, statistically significant treatment effects were found compared with expected physiological progression (p < 0.05).

## DISCUSSION

This retrospective, multicenter, real-world study demonstrates the efficacy of HAL lenses in slowing the progression of myopia. At 12 months post-HAL wear, myopia progression and axial elongation slowed significantly compared to the untreated period and trajectory-adjusted expected physiological progression (p = 0.001). HAL lenses showed significant slowing of myopia progression in both age groups (≤10 years and >10 years). The slowing of myopia progression with HAL was significant in both groups, those receiving atropine and those without atropine, relative to expected physiological progression. This study, to the best of the authors’ knowledge, is the first to provide evidence from Indian children and adolescents regarding the influence of age and adjunctive atropine on the efficacy of HAL lens treatment.

Published age-adjusted meta-regression models,[26,27] were used as a reference to compare treatment outcomes with trajectory-adjusted expected physiological progression. This comparison enabled the interpretation of our findings. Previous literature reported faster myopia progression in Asian children, accounting for greater progression of SER (42.2% higher)[26] and axial length (41% higher)[27]. Since the present study included Indian children and adolescents, it highlights the need for region-specific data to better understand progression patterns and treatment responses in the Indian population.

In this study, the HAL lens successfully slowed the progression of myopia, which is in accordance with previous studies. Post 1-year of HAL lens wear, myopia progression slowed (mean SER change: −0.72 D in the untreated year and −0.11 D during 1 year with HAL; mean axial elongation: 0.11 mm with HAL over 1 year) compared to the untreated year. Similarly, Bao et al. in a 2-year study reported the HAL group with lesser SER progression by a mean (SE) of 0.80±0.11 compared to the SVL group with a mean SER progression of 1.46 D.[22] Additionally, in the HAL group, the axial elongation was reduced by 0.35 ±0.05 mm [22], comparable to the present study, where axial elongation was 0.11±0.04 mm during HAL lens wear. Li et al. investigated the 5-year myopia progression and reported that the HAL lens slowed myopia progression by 1.75 D and axial elongation by 0.72 mm.[25] In another Indian prospective study including children aged 5 to 15 years, at the 1-year follow-up, the rate of myopia progression was reduced in the HAL group(0.36 ± 0.12 D/year).[28] The magnitude and direction of effect observed with the HAL lens in slowing of myopia progression among the Indian population align with international studies.

In our cohort, children receiving atropine in combination with HAL lenses showed slightly slower myopia progression and axial elongation. This finding is consistent with published literature that has highlighted the efficacy of low-dose atropine in slowing myopia progression among Indian children.[29–32] Hu et al. found that combining HAL and 0.01% atropine slowed progression by 0.338 D compared with atropine alone (p=0.0001) in children with SER > –2.00 D.[33] Similarly, Sim et al. reported that adding HAL to low-dose atropine reduced progression from –0.60 D and 0.24 mm over six months to –0.15 D and 0.14 mm over 12 months in children already progressing on atropine alone.[34] While HAL lenses alone provide substantial control, the addition of atropine may enhance efficacy in children with a higher risk of myopia progression or inadequate response. In the present study, both HAL monotherapy and HAL combined with atropine demonstrated statistically significant slowing relative to expected physiological progression.

The SOLIDITY study is one of the largest real-world investigations on the effectiveness of HAL lenses, conducted across 10 ophthalmic centres in India. The multicentre design allowed generalization of the findings into routine clinical practice, thereby improving patient care. The subgroup analyses by age showed the real-world effectiveness of HAL in older children, highlighting clinically meaningful differences depending on age groups and concomitant use of atropine. These findings were consistent with trajectory-adjusted modelling of expected physiological progression. As the first real-world study in India to evaluate the use of HAL (Essilor^®^ Stellest^®^) lenses, the SOLIDITY study offers valuable insights to inform clinical adoption of HAL lenses across diverse populations

Several limitations of the study must be acknowledged. The retrospective design introduces the potential for selection bias and uncontrolled confounding. Additional factors known to influence myopia, such as outdoor exposure, near-work activity, and socioeconomic background, were not accounted for in the present study. Axial length values from the untreated year were not available; therefore, comparison was limited to the period during which participants were wearing HAL lenses. Participants with biologically implausible axial length reductions (>0.1 mm) were excluded, and those with minor axial length reductions (≤0.1 mm) were included. The follow-up interval was standardized to visits at 12 ± 1 months, which involved adjustment of outlier timepoints to a 12-month equivalent. Such adjustments would possibly introduce minor inaccuracies. However, trajectory-adjusted modelling and multiple imputation were used to minimize potential bias arising from variable follow-up intervals. The retrospective design limited standardization of refraction and biometry instruments across centers; however, all measurements were obtained under cycloplegia using validated clinical devices in specialized myopia clinics. Furthermore, exclusion of one child with myopia progression >–2.0 D/year at baseline, and 17 with SER >–8.0 D at baseline, limits the generalization to children and adolescents with rapid myopia progression and high myopia. The present study enrolled 372 children, of whom 221 had a follow-up duration of ≥12 months, while 151 had a follow-up period of 6 to 12 months. These findings underscore the need for long-term, prospective studies to comprehensively evaluate outcomes in the Indian pediatric population.

## CONCLUSION

The present study demonstrates the real-world evidence validating the use of HAL lenses as an effective myopia control intervention in Indian children and adolescents, with statistically significant slowing observed relative to trajectory-adjusted expected physiological progression. HAL lenses constitute a simple, practical, and non-invasive treatment modality for mitigating myopia progression in the paediatric population in India. Future prospective, long-term studies are warranted to establish the clinical significance of treatment effects and determine optimal combination regimens for sustained myopia control.

## Data Availability

The data supporting the findings of this study are available from the corresponding author upon reasonable request, subject to compliance with applicable privacy and ethical considerations.

## Acknowledgments

We thank all the participating ophthalmic centres involved in the SOLIDITY study for their collaboration and contribution to data collection. We thank Mark Bullimore for his comments on previous versions of the manuscript; Dr. Gunjan Chauhan for her medical writing and editorial assistance; and Mitravinda for assistance with statistical analysis.

## Supporting information captions

Table S1: Comparison of unimputed and imputed values for spherical equivalent refraction (SER) and axial length (AL)

*a* Unimputed dataset (complete-case analysis), n = 217.

*b* Full dataset after multiple imputation using Multiple Imputation by Chained Equations (MICE), n = 372.

SER: spherical equivalent refraction; AL: axial length; SD: standard deviation; T1: untreated year (one year prior to HAL prescription); T2: baseline year (HAL prescription); T3: treated year (one year post-HAL wear.

Missing and out-of-window values were imputed using MICE to preserve the multivariable structure of the data and generate physiologically plausible 12-month equivalent values.

Comparisons between unimputed and imputed datasets were performed using independent-samples t-tests. Non-significant differences between unimputed and imputed datasets confirm that imputation did not introduce systematic bias and preserved the underlying distribution of SER and AL measurements.

Figure S1: Subject Selection and Data Inclusion Criteria for the ‘SOLIDITY’ Study. Flow diagram showing subject screening, exclusion criteria, and final analysis population. Of 514 subjects screened across 10 ophthalmic centres in India, 372 met the eligibility criteria and were included in the final analysis. Subjects were excluded due to missing baseline spherical equivalent refraction (SER) values, protocol violations, biologically implausible axial length (AL) or SER progression values, prior myopia control interventions, duplicate records, or unverifiable source data. Multiple imputation by chained equations (MICE) was applied to account for missing or out-of-window follow-up measurements, resulting in a full analysis dataset of 372 subjects, including 217 complete cases and 155 imputed cases. The final analysis population was used for trajectory-adjusted modelling of SER progression and age-adjusted modelling of axial elongation based on published meta-regression models.

## Notes

### Competing Interest Statement

KS and OP are employees of Essilor Luxottica. MB serves as a consultant and scientific advisor to Essilor Luxottica and has received consultancy fees and/or honoraria for advisory activities. The remaining authors declare no competing interests.

### Funding Statement

Yes

### Author Declarations

Ethical approval was obtained from the ACE Independent Ethics Committee and subsequently from institutional review boards at all participating centres. Due to the retrospective and anonymised nature of the data, individual informed consent was waived.

